# Impact of ethnicity on outcome of severe COVID-19 infection. Data from an ethnically diverse UK tertiary centre

**DOI:** 10.1101/2020.05.02.20078642

**Authors:** James TH Teo, Daniel M Bean, Rebecca Bendayan, Richard JB Dobson, Ajay M Shah

**Affiliations:** King’s College Hospital NHS Foundation Trust, London, U.K.; Department of Clinical Neuroscience, Institute of Psychiatry, Psychology and Neuroscience, King’s College London, London, U.K.; Department of Biostatistics and Health Informatics, Institute of Psychiatry, Psychology and Neuroscience, King’s College London, London, U.K.; Health Data Research UK London, Institute of Health Informatics, University College London, London, U.K.; NIHR Biomedical Research Centre at South London and Maudsley NHS Foundation Trust and King’s College London, London, U.K.; School of Cardiovascular Medicine & Sciences, King’s College London British Heart Foundation Centre of Excellence, London, U.K.

## Abstract

During the current COVID-19 pandemic, it has been suggested that BAME background patients may be disproportionately affected compared to White but few detailed data are available. We took advantage of near real-time hospital data access and analysis pipelines to look at the impact of ethnicity in 1200 consecutive patients admitted between 1st March 2020 and 12th May 2020 to King’s College Hospital NHS Trust in London (UK).

Our key findings are firstly that BAME patients are significantly younger and have different co-morbidity profiles than White individuals. Secondly, there is no significant independent effect of ethnicity on severe outcomes (death or ITU admission) within 14-days of symptom onset, after adjustment for age, sex and comorbidities.

## Introduction

As the Covid-19 pandemic extends from ethnically homogeneous countries in East Asia to multicultural populations in Europe and North America, anecdotal reports suggest a higher disease burden in Black, Asian and Minority Ethnic (BAME) groups^1^. Early UK audit data on COVID-19 indicate a higher proportion of BAME patients on critical care units than previous years but do not establish whether disease outcome is worse than in White patients or the factors underlying any disparity^2^.

## Methods

This project operated under London South East Research Ethics Committee approval (reference 18/LO/2048) granted to the King’s Electronic Records Research Interface (KERRI). Specific work on COVID19 research was reviewed with expert patient input on a virtual committee with Caldicott Guardian oversight.

We evaluated the effect of ethnicity on disease outcomes in a consecutive cohort of 1200 patients admitted with COVID-19 disease to King’s College Hospital NHS Foundation Trust from 1st March 2020 to 12th May 2020. This multi-site tertiary centre in South East London serves an ethnically diverse population: approximately 30% of residents identify as BAME in the immediate vicinity of the largest site (boroughs of Southwark and Lambeth) and ~12% near the Bromley site^3^.

We used an informatics pipeline previously described for evaluating ACE-inhibitor risk in patients with COVID-19 disease^4^. The primary endpoint was death (WHO-COVID-19 Outcomes Scale 8) or admission to critical care (WHO-COVID-19 Outcomes Scale 6-7) by 14 days after symptom onset. Self-assigned ethnicity is reported according to UK census categories.

## Results

The distribution of self-identified ethnicity among patients is shown in Table 1. 30.7% of patients were BAME. BAME patients (84% Black) had a significantly higher prevalence of diabetes and hypertension but were significantly younger than White patients (Table 2, Figure 1). The prevalence of ischaemic heart disease (IHD) and heart failure tended to be lower in BAME compared to White patients. The incidence of the primary end-point by 21 days after symptom onset and Kaplan-Meier curves were similar between White and BAME patients (Figure 2).

**Table 1.** Clinical characteristics of study cohort. The “Unknown/Mixed/Other Ethnicity” group contains any single ethnicity that is not White or BAME, any mixed ethnicity, and any missing / not stated ethnicity. BAME = Black, Asian and Minority Ethnic. All values are N (%) except age which is mean (s.d.). All percentages are relative to column total except for the total N row, which shows percent of cohort.

|  | Overall | BAME<br>Ethnicity | Black<br>Ethnicity | White<br>Ethnicity | Asian<br>Ethnicity | Unknown/<br>Mixed/Other<br>Ethnicity |
| --- | --- | --- | --- | --- | --- | --- |
| N (%) | 1200 | 368 (30.7) | 310 (25.8) | 512 (42.7) | 58 (4.8) | 320 (26.7) |
| Age | 68.0 (17.1) | 62.9 (16.4) | 63.0 (16.1) | 73.7 (15.6) | 62.3 (18.2) | 64.7 (17.4) |
| Male | 686 (57.2%) | 212 (57.6%) | 172<br>(55.5%) | 291 (56.8%) | 40 (69.0%) | 183 (57.2%) |
| Hypertension | 645 (53.8%) | 233 (63.3%) | 201<br>(64.8%) | 246 (48.0%) | 32 (55.2%) | 166 (51.9%) |
| Diabetes<br>mellitus | 418 (34.8%) | 179 (48.6%) | 152<br>(49.0%) | 126 (24.6%) | 27 (46.6%) | 113 (35.3%) |
| Ischaemic<br>heart disease<br>or heart failure | 224 (18.7%) | 54 (14.7%) | 43 (13.9%) | 109 (21.3%) | 11 (19.0%) | 61 (19.1%) |
| Primary<br>Endpoint<br>(Death or<br>Critical Care) | 415 (34.6%) | 123 (33.4%) | 94 (30.3%) | 185 (36.1%) | 29 (50.0%) | 107 (33.4%) |
| Death | 288 (24.0%) | 68 (18.5%) | 53 (17.1%) | 151 (29.5%) | 15 (25.9%) | 69 (21.6%) |
| Critical Care | 168 (14.0%) | 65 (17.7%) | 49 (15.8%) | 51 (10.0%) | 16 (27.6%) | 52 (16.2%) |

**Table 2.** Statistical tests for differences in patient characteristics. All tests compare BAME ethnicity vs White ethnicity. Continuous variables are compared using a t-test, count variables are compared using a Chi-squared test. P (corrected) is the p-value after Bonferroni correction for multiple comparisons.

| Variable | Test | p | p (corrected) |
| --- | --- | --- | --- |
| Age | t-test | <0.001 | <0.001 |
| Male | Chi2 | 0.87 | 1 |
| Hypertension | Chi2 | <0.001 | <0.001 |
| Diabetes | Chi2 | <0.001 | <0.001 |
| Ischaemic heart disease or heart failure | Chi2 | 0.016 | 0.097 |
| Endpoint Status | Chi2 | 0.45 | 1 |

**Figure 1.**
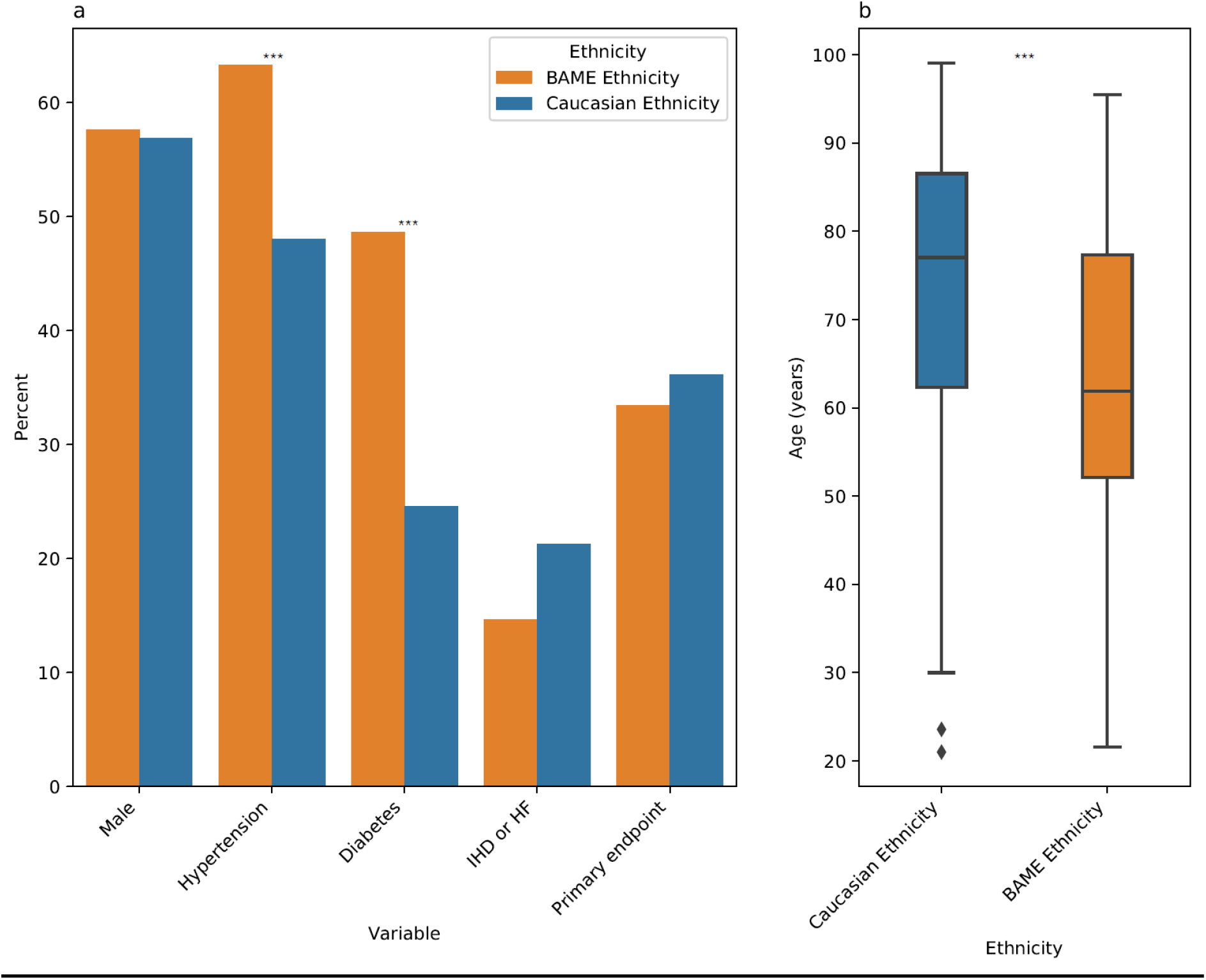
Key demographics by Ethnicity. a) Rate of male sex, hypertension, diabetes, ischaemic heart disease or heart failure and primary endpoint. b) Distribution of age in years. BAME = Black, Asian and Minority Ethnic, IHD = ischaemic heart disease, HF = heart failure. Asterisks indicate significance; * = p < 0.05, ** = p < 0.01, *** = p < 0.001, all others not significant (p ≥ 0.05).

**Figure 2:**
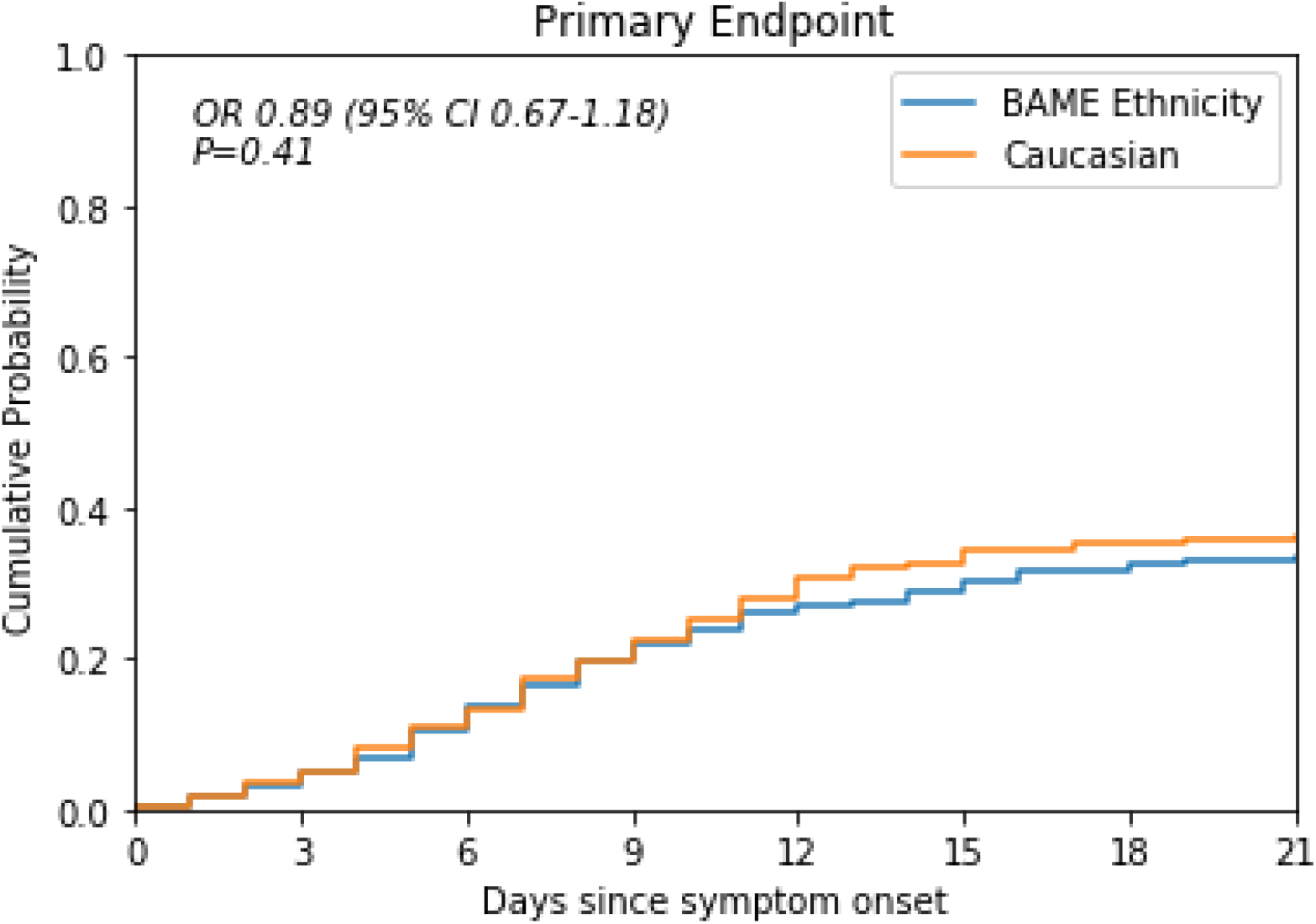
Kaplan-Meier survival curves for patients of BAME ethnicity (blue) or White ethnicity (orange). Note “survival” here indicates “did not reach primary end-point”.

We tested whether BAME ethnicity was independently associated with the incidence of the primary outcome but did not find a significant statistical association in a series of multivariate logistic regression models adjusted for age, sex and comorbidities (hypertension, diabetes, ischaemic heart disease/heart failure) - Table 3. Increasing age and male sex were associated with an increased likelihood of the primary outcome.

**Table 3.** Summary of odds ratios in statistical models. Model 1 is univariate, Models 2 and 3 are multivariate. Odds ratios and p-values calculated from logistic regressions applying Firth’s correction^5^. BAME = Black, Asian and Minority Ethnic Groups.

| Variable | OR (95% CI) | P-value | Model |
| --- | --- | --- | --- |
| BAME Ethnicity | 0.89 (0.67-1.18) | 0.41 | Model 1 |
| BAME Ethnicity | 1.09 (0.81-1.47) | 0.58 | Model 2 |
| Age per 10 years | 1.23 (1.12-1.34) | <0.001 |  |
| Male | 1.49 (1.12-1.99) | <0.01 |  |
| BAME Ethnicity | 1.11 (0.80-1.52) | 0.53 | Model 3 |
| Age per 10 years | 1.25 (1.14-1.38) | <0.001 |  |
| Male | 1.50 (1.12-2.00) | <0.01 |  |
| Hypertension | 0.91 (0.66-1.24) | 0.54 |  |
| Diabetes | 1.02 (0.74-1.40) | 0.91 |  |
| Ischaemic heart disease or heart failure | 0.76 (0.52-1.10) | 0.15 |  |

This study in a series of 1200 patients hospitalised with COVID-19 disease in London (United Kingdom) indicates large ethnicity-related differences in age and comorbidities. Patients of BAME origin who require admission for COVID-19 are significantly younger than White patients from the same geographical area and have a significant higher burden of diabetes and hypertension. We did not identify an independent impact of ethnicity on the severity of in-hospital outcome after admission. Our BAME patients were predominantly Black and the results may not apply to other ethnicities. Further studies are required to establish the reasons underlying the ethnicity-related differences in number and profile of patients who require hospitalisation for COVID-19 disease.

## Data Availability

Confidential patient data are not available in accordance with Data Confidentiality Regulations

## Acknowledgements

DMB is funded by a UKRI Innovation Fellowship as part of Health Data Research UK MR/S00310X/1 (https://www.hdruk.ac.uk). RB is funded in part by grant MR/R016372/1 for the King’s College London MRC Skills Development Fellowship programme funded by the UK Medical Research Council (MRC, https://mrc.ukri.org) and by grant IS-BRC-1215-20018 for the National Institute for Health Research (NIHR, https://www.nihr.ac.uk) Biomedical Research Centre at South London and Maudsley NHS Foundation Trust and King’s College London. RJBD is supported by Health Data Research UK; The BigData@Heart Consortium, European Union’s Horizon 2020 grant agreement No. 116074; The National Institute for Health Research (NIHR) University College London Hospitals Biomedical Research Centre and the NIHR Biomedical Research Centre at South London and Maudsley NHS Foundation Trust and King’s College London; and the UK Research and Innovation London Medical Imaging & Artificial Intelligence Centre for Value Based Healthcare. AMS is supported by the British Heart Foundation (CH/1999001/11735), the NIHR Biomedical Research Centre at Guy’s & St Thomas’ NHS Foundation Trust and King’s College London (IS-BRC-1215-20006), and the Fondation Leducq. The views expressed are those of the author(s) and not necessarily those of the NHS, the NIHR or the Department of Health and Social Care.

We thank all the clinicians managing the patients, the patient experts of the KERRI committee, Professor Clive Kay, Professor Irene Higginson, Professor Alastair Baker and Professor Jules Wendon for their support.

